# Adverse events following COVID-19 virus vaccination in Japanese young population: The first cross-sectional study conducted by a questionnaire survey after the first-time-injection

**DOI:** 10.1101/2021.07.23.21261029

**Authors:** Marie Suehiro, Shinya Okubo, Kensuke Nakajima, Kosuke Kanda, Masanobu Hayakawa, Shigeru Oiso, Tsutomu Kabashima, Hideaki Fujita, Yukio Ando, Takahiro Muro

## Abstract

A cross-sectional study was conducted to clarify the adverse events of COVID-19 vaccines in Japanese young population.

The proportion of participants with adverse events at the vaccination site (immediately or within 30 minutes after vaccination) was 0.5%, and anaphylaxis occurred in one female student (0.03%).

We analyzed 1,877 data obtained from a questionnaire survey of 1,993 vaccinated individuals. Eighty-two percent of participants complained of local adverse events. Injection site pain was the most common local adverse event (71%). Systemic adverse events occurred in 48% of participants. The most common adverse event was myalgia (34%).

A multivariable logistic regression model was used to determine risk factors.

Local adverse events were associated with sex (female) and allergy history, with odds ratios (ORs) (95% confidence interval [CI]) of 2.15 (1.69-2.73) and 1.73 (1.10-2.74), respectively. Systemic adverse events were associated with sex (female), age (<20 years old), allergy history, and history of adverse events with previous medications, with ORs (95% CI) of 2.49 (2.03-3.06), 1.80 (1.44-2.24), 1.39 (1.03-1.89), and 1.53 (1.02-2.29).

The results of this study clarified for the first time that age less than 20 years is a risk factor for systemic adverse events from the COVID-19 Vaccine Moderna Intramuscular Injection. This information will give impacts on considering adverse events and its mechanisms in mRNA vaccination.

## Introduction

The coronavirus disease 2019 (COVID-19) crisis has been spreading worldwide. The COVID-19 vaccine is expected to be an effective preventive measure.

In Japan, vaccinations for priority groups, such as medical staff and elderly people (older than 65 years old), were being conducted first. Vaccinations at workplaces began on June 21, 2021 in limited universities and companies, and our university employees and students were subject to vaccination.

Currently, in Japan, the COMIRNATY intramuscular injection is used for priority vaccinations, and COVID-19 Vaccine Moderna Intramuscular Injection is used for vaccination in workplaces.

The adverse events of the coronavirus vaccine in elderly people have been studied; however, those of the coronavirus vaccine in young people have not been studied in Japan.

The reported adverse events were following vaccination using the COMIRNATY intramuscular injection, and there have been no reports of adverse events following the COVID-19 Vaccine Moderna Intramuscular Injection in Japanese population ^1,2)^.

In addition, racial differences in the adverse events following coronavirus vaccines have not been sufficiently studied.

Therefore, to clarify the adverse events following coronavirus vaccination in young population, we completed a questionnaire survey after the first dose for students, faculty, and staff who belong to an educational foundation, Kyushu Bunka Gakuen.

## Study population and method

### Design

This study used a cross-sectional design with a questionnaire survey.

### Study population

The study population included people who had a first time vaccination using COVID-19 Vaccine Moderna Intramuscular Injection on workplace vaccination of an educational foundation, Kyushu Bunka Gakuen.

This workplace vaccination included students attending educational facilities run by the educational foundation Kyushu Bunka Gakuen, universities, junior colleges, training facilities for cooking, training facilities for dental hygienists, and high schools.

In addition, this workplace vaccination included those employed by local companies related to the educational foundation of Kyushu Bunka Gakuen.

### Data collection

Information regarding adverse events at the vaccination site was obtained from medical records.

To obtain information on that adverse events occurred after leaving the site, we conducted a questionnaire survey on the website.

An explanation of this study was provided on the website, and answers to questions regarding consent to research were obtained one by one.

Following adverse events related to vaccination, we obtained the following in a questionnaire: type of adverse event, date of occurrence, period of occurrence, and medication/care.

In addition, the attributes (age, sex, allergy history, and history of adverse events with previous medication) of the respondents were also obtained. Fever was defined as an increase of more than 1°C above the usual body temperature.

### Statistical analysis

Patients’ demographics and answers to the questionnaire are summarized with frequencies and percentages for categorical data and median plus range for continuous data.

We compared the patients who experienced adverse events (AE) and those who did not experience adverse events (No-AE) using the Wilcoxon rank sum test for continuous variables and Fisher’s exact test for dichotomous variables. Differences were considered significant at p < 0.05.

To estimate the risk factors of adverse events, we first evaluated the odds ratio (OR), 95% confidence intervals (95% CI), and p-values of each potential risk factor using unadjusted logistic regression models.

Second, we determined the risk factors using a multivariable logistic regression model. Statistical significance was determined by 95% CIs not including 1.00 in logistic analyses. Candidate predictors were selected according to a literature review and clinical expertise. We selected four variables, including patient demographics (age, sex, allergy history, and history of adverse events for past medication). Age, a continuous variable, was categorized as minor (<20 years old) or adult.

All statistical analyses were performed using JMP Pro 16 (SAS Institute Inc., Cary, NC, USA).

### Ethics statement

This study was approved by the Nagasaki International University Ethics Committee (Approval number 50, July 2, 2021).

## Results

### Adverse events at the vaccination site

In this workplace vaccination program, 3,998 people were vaccinated.

Adverse events occurring at the vaccination site (immediately or within 30 min after vaccination) are shown in Table 1.

**Table 1.** Adverse events occurring at the vaccination site

(immediately or within 30 min after vaccination)
| Adverse events | Number of<br>people | (%) | sex |  |
| --- | --- | --- | --- | --- |
|  |  |  | female | male |
| Vagal syncope | 5 | 0.13 | 4 | 1 |
| Hyperventilation syndrome | 4 | 0.10 | 4 |  |
| Feeling sick | 4 | 0.10 | 3 | 1 |
| Suffocation | 3 | 0.08 | 3 |  |
| Nausea | 1 | 0.03 | 1 |  |
| Anaphylaxis | 1 | 0.03 | 1 |  |
| Hyperventilation with involuntary movement | 1 | 0.03 | 1 |  |
| Severe urticaria | 1 | 0.03 | 1 |  |
| Total | 20 | 0.5 | 18 | 2 |

The proportion of individuals with adverse events at the vaccination site was 0.5%, and anaphylaxis occurred in one female student who had anaphylaxis at influenza vaccination once (0.03%).

### Demographic characteristics of participants

Table 2 shows the demographic characteristics of participants in the questionnaire survey. We obtained data from 1,993 subjects who responded to the questionnaire survey (response rate 49.8%). A total of 1,877 subjects who agreed to participate in the present study were enrolled in the analysis. Female participants accounted for 66% of the study population, with a median age of 22 years. Twenty-four percent of the study population was younger than 20 years old.

**Table 2.** Demographic Characteristics of participants

| Characteristic | Number of people<br>(N=1877) | (%) <sup>a)</sup> |
| --- | --- | --- |
| Female | 1242 | 66 |
| Age, median (range), y | 22 | (18-70) |
| Age (minor*) | 444 | 24 |
| Allergy history | 219 | 12 |
| Adverse events history for past medication | 118 | 6 |

a) Data are expressed as No.(%) unless otherwise indicated.
\*minor : <20 years old

### Adverse events after leaving the vaccination site

The incidence of each adverse event is presented in Table 3. Local and systemic adverse events were generally mild. Eighty-two percent of participants complained of local adverse events. Injection site pain was the most common local adverse event (71%). Systemic adverse events occurred in 48% of participants. The most common adverse event was myalgia (34%), followed by general fatigue (31%).

**Table 3.** The incidence of each adverse event

| Adverse events | Number of people | (%) |
| --- | --- | --- |
| Local adverse events |  |  |
| Any | 1533 | 82 |
| Pain | 1334 | 71 |
| Swelling | 757 | 40 |
| thermal sense | 641 | 34 |
| Redness | 476 | 25 |
| Itching | 370 | 20 |
| Induration | 312 | 17 |
| Systemic adverse events |  |  |
| Any | 894 | 48 |
| Myalgia | 632 | 34 |
| Fatigue | 581 | 31 |
| Fever | 416 | 22 |
| Headache | 386 | 21 |
| Joint pain | 119 | 6 |
| Chills | 86 | 5 |

Figure 1 shows the time of onset of adverse events, Figure 2 disclosed the duration of adverse events, and the type of treatment for adverse events are demonstrated in Figure 3. Injection site pain occurred from day 0 to day 1 and continued for a couple of days. However, almost all the symptoms improved without any treatments. The percentage of participants with itching and local redness after day 7 were 27% and 19%, respectively. The onset of myalgia was often on the day and the day after injection (98%), and symptoms lasted 2-3 days, which was a little longer than other systemic adverse reactions (73%, 31% for 3 days). Of those who developed symptoms, 75% did not undergo any treatments, and 17% were administered acetaminophen.

**Fig. 1.**
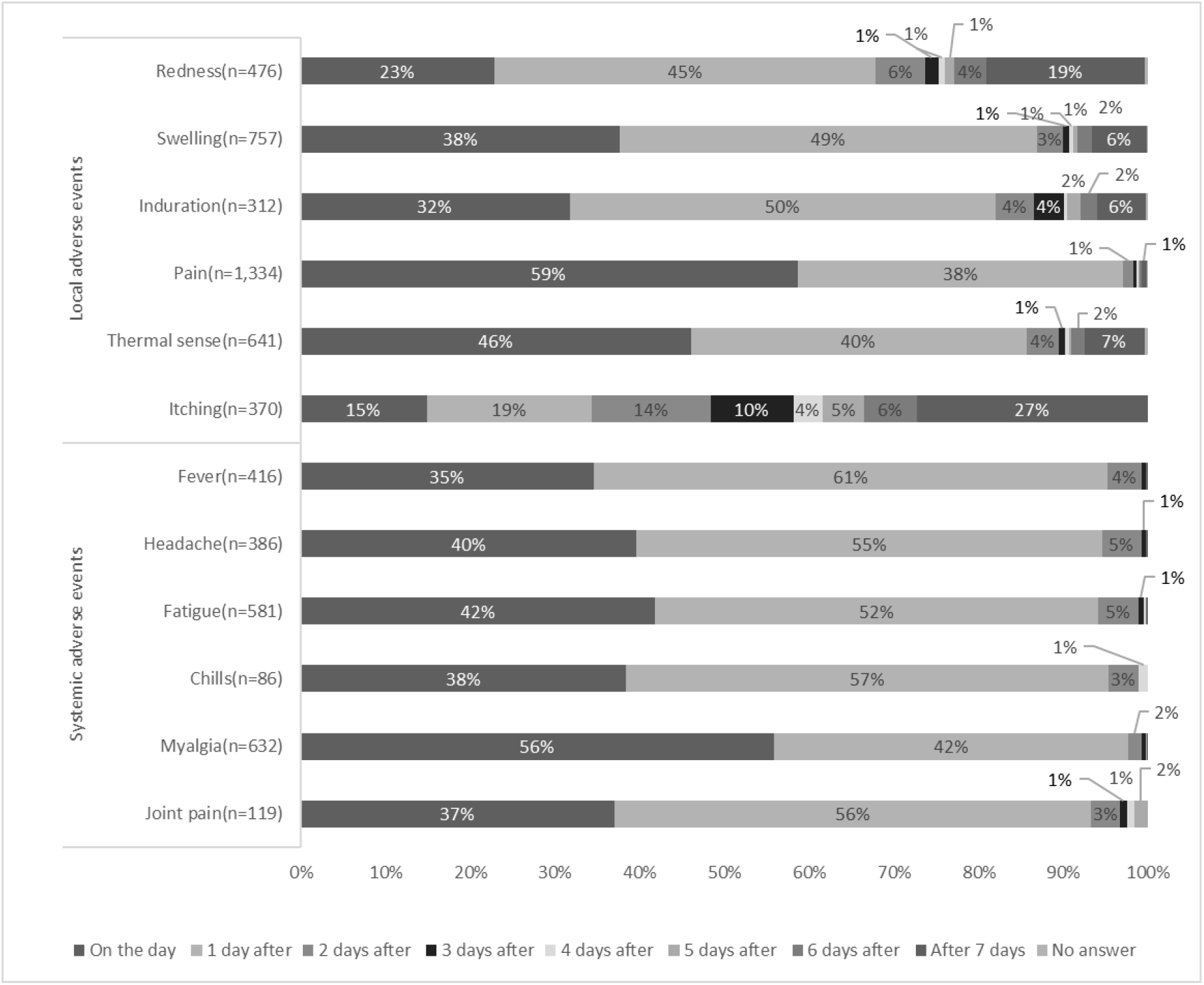
The time of onset of adverse events (days after vaccination)

**Fig. 2.**
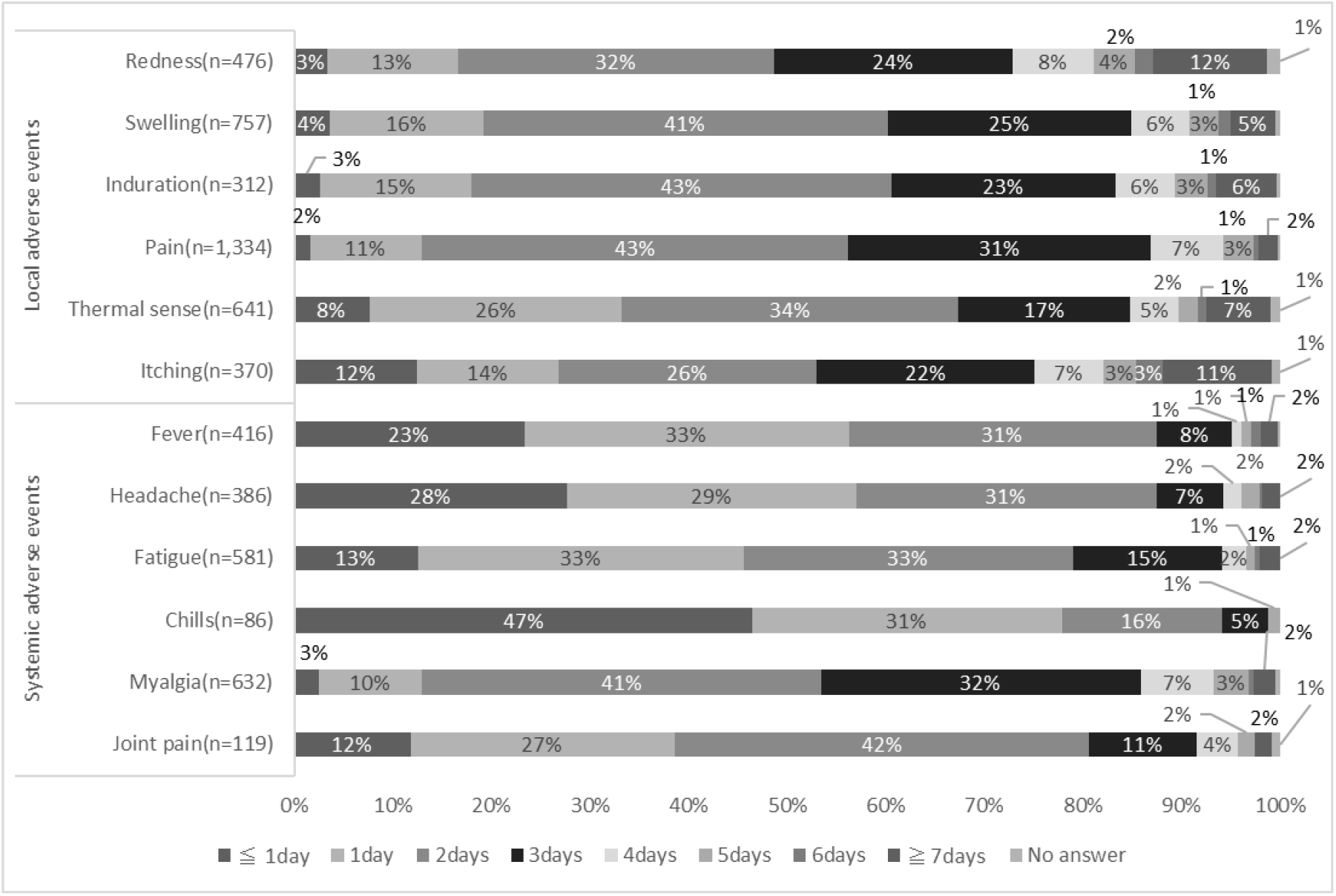
The duration of adverse events.

**Fig. 3.**
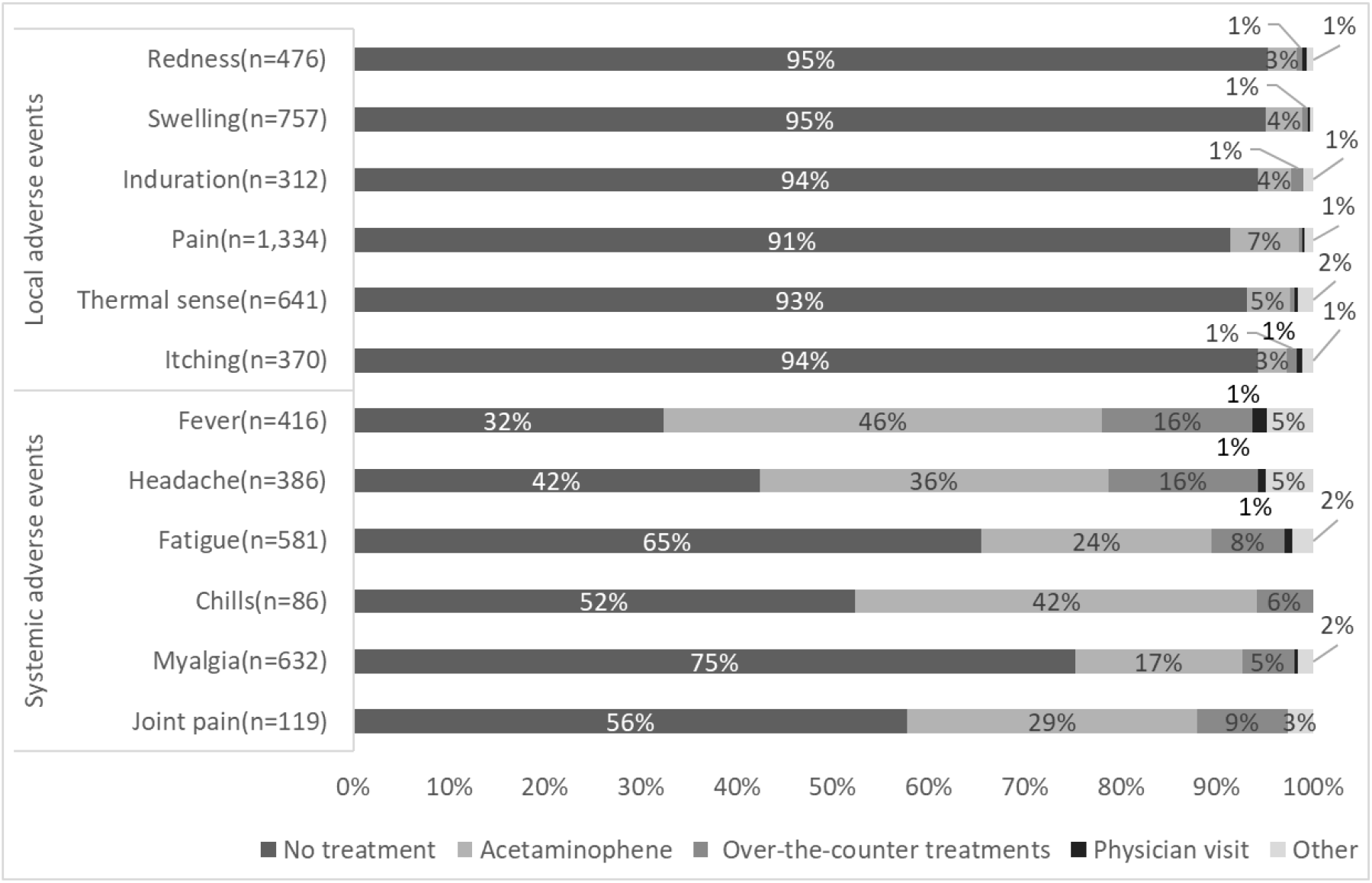
The type of treatment for adverse events.

General fatigue often occurred the day of injection (94%) and continued for 2-3 days thereafter (66%).

### Comparison of the AE and No-AE groups

The characteristics of the AE and No-AE groups are shown in Table 4. Regarding local adverse events, the number of female participants in the AE group was significantly larger than that in the No-AE group (p < 0.001). Similarly, the number of people with an allergy history of AE was greater than that of No-AE (p= 0.003). Regarding systemic adverse events, the number of female and minors in the AE group was significantly higher than the number in the No-AE group (p < 0.001). The number of people with an allergy history in the AE group was greater than that in the No-AE group (p = 0.004).

**Table 4.** The characteristics of the AE and No-AE groups^a)^

**Local adverse events**
| Characteristic | AE<sup>b)</sup><br>(N=1,533) | No-AE<sup>b)</sup><br>(N=344) | P-Value<sup>c)</sup> |
| --- | --- | --- | --- |
| sex (female) | 1068 (70%) | 174 (51%) | <0.001 |
| age, median,(range),y | 22 (18-70) | 26 (18-64) | <0.001 |
| age (minor)<sup>d)</sup> | 374 (24%) | 70 (20%) | 0.139 |
| allergy history | 194 (17%) | 25 (7%) | 0.003 |
| adverse events history for past medication | 103 (7%) | 15 (4%) | 0.006 |

**Systemic adverse events**
| Characteristic | AE<sup>b)</sup><br>(N=894) | No-AE<sup>b)</sup><br>(N=983) | P-Value<sup>c)</sup> |
| --- | --- | --- | --- |
| sex (female) | 691 (77%) | 551 (56%) | <0.001 |
| age,median,(range),y | 21 (18-70) | 27 (18-64) | <0.001 |
| age (minor)<sup>d)</sup> | 267 (30%) | 177 (18%) | <0.001 |
| allergy history | 125 (14%) | 94 (10 %) | 0.004 |
| adverse events history for past medication | 71 (8%) | 47 (5%) | 0.111 |

a) Data are expressed as No.(%) unless otherwise indicated.
b)Abbreviations:AE, the patients who experienced adverse events; No-AE, the patients who did not experience adverse events
c) Wilcoxon's rank sum test was performed for continuous variables, and Fisher's exact test was performed for categorical variables.
d) minor : <20 years old

### Multivariate analysis of AE and No-AE groups

Results of the multivariable analysis of the AE and No-AE groups are shown in Table 5. Local adverse events were associated with sex (female) and allergy history, with ORs (95% CI) of 2.15 (1.69-2.73) and 1.73 (1.10-2.74), respectively. Looking in detail at individual adverse events, in local adverse events, redness and thermal sense showed similar tendencies. Induration and itching were associated with age (<20 years old) (OR [95% CI]: 0.56 [0.40-0.78] and 0.62 [0.46-0.83], respectively), and injection site pain was associated with a history of adverse events with prior medication OR [95% CI]: 3.11 [1.12-8.65]).

**Table 5.**
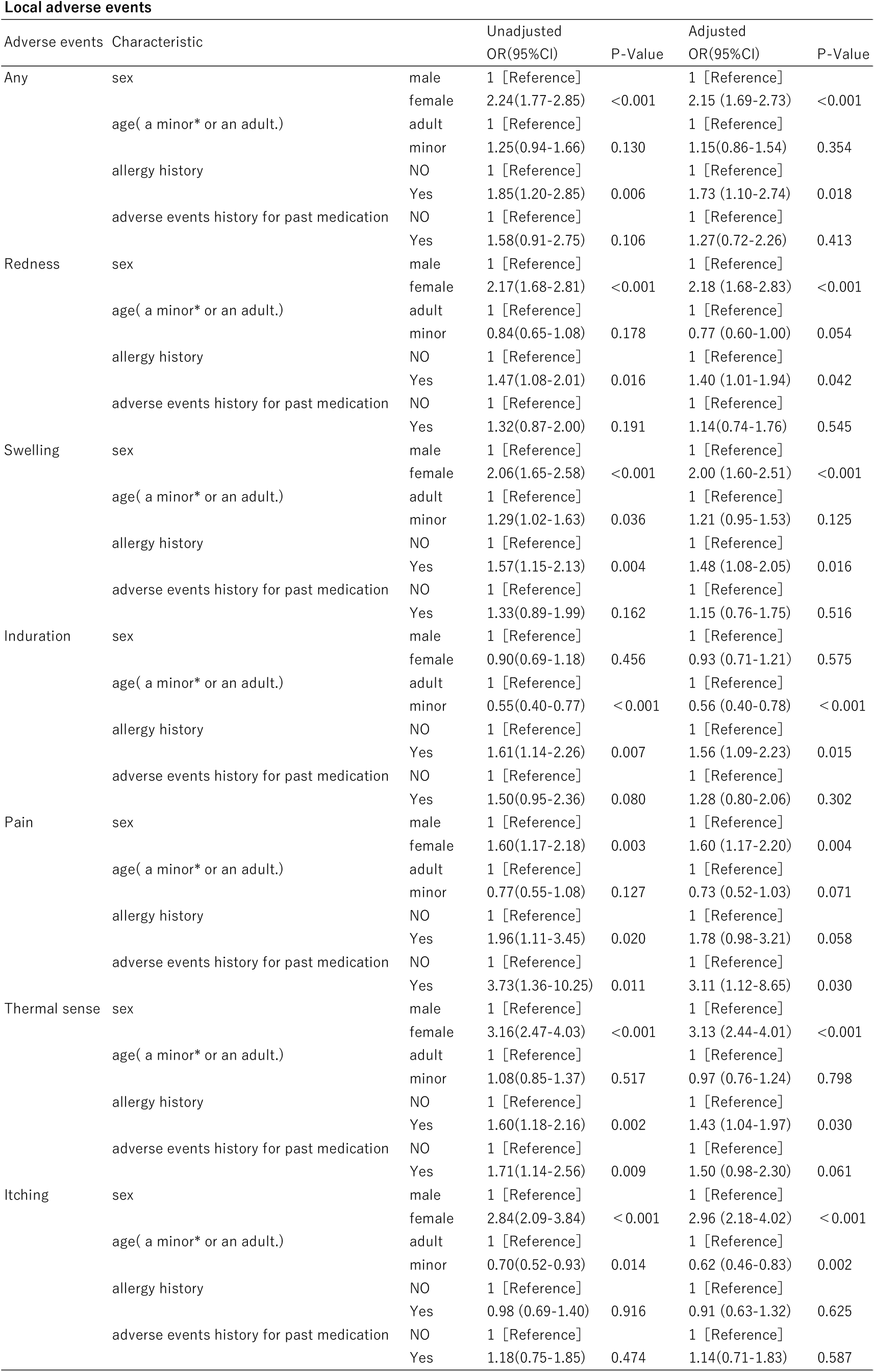
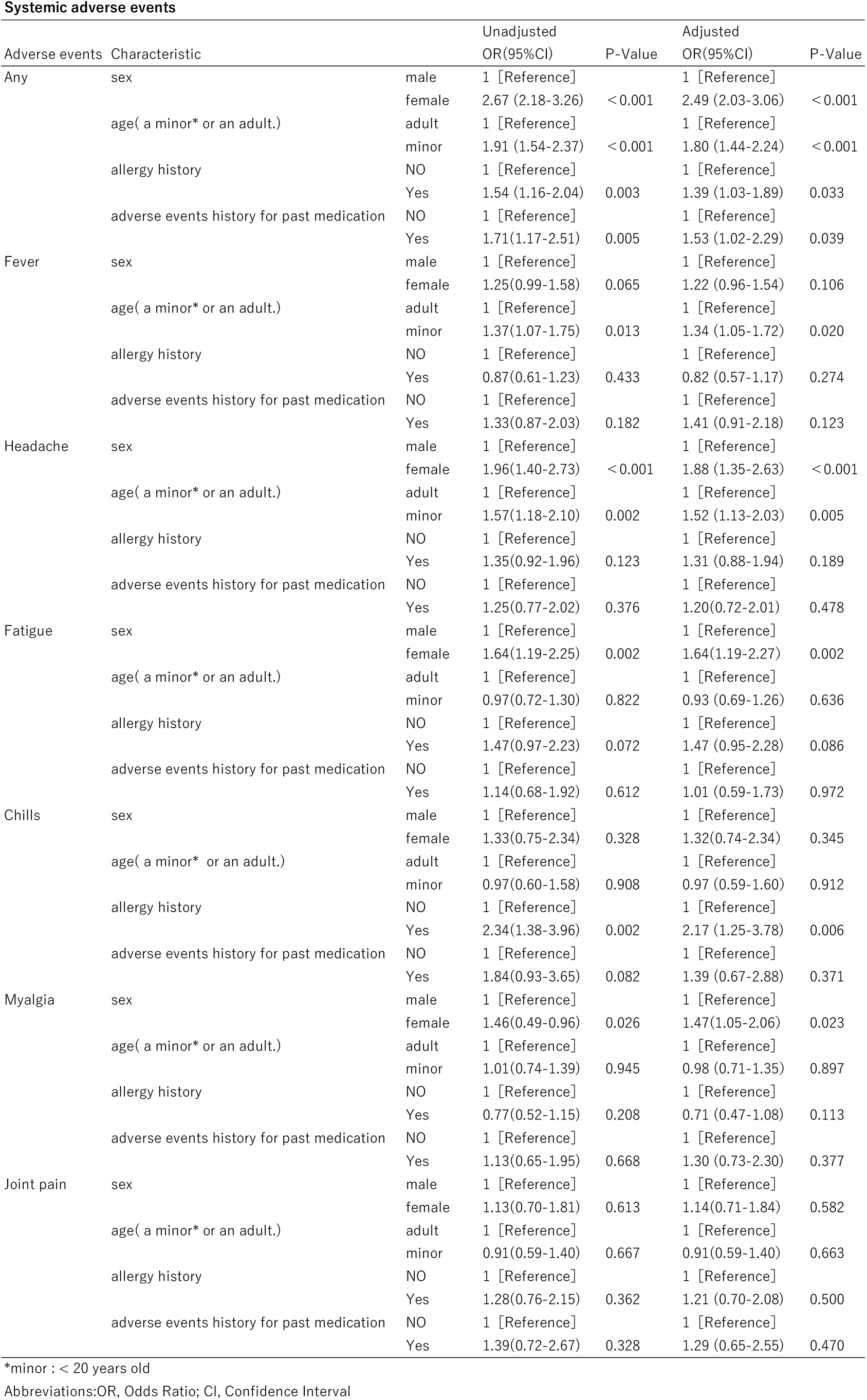
Results of the multivariable analysis of the AE and No-AE groups

Systemic adverse events were associated with sex (female), age (<20 years old), allergy history, and history of adverse events with past medications, with ORs (95% CI) of 2.49 (2.03-3.06), 1.80 (1.44-2.24), 1.39 (1.03-1.89), and 1.53 (1.02-2.29), respectively.

## Discussion

The results of this study clarified for the first time that minors aged <20 years old are at greater risk of systemic adverse events from the COVID-19 Vaccine Moderna Intramuscular Injection.

According to a report by the Japanese Self-Defense Forces, the incidences tended to be higher in younger people; the factors analysed in this study confirmed this statistically^1)^.

Second, the results of this study showed that female sex is a major risk factor for many adverse events.

According to Ministry of Health, Labour, and Welfare, Japan and the CDC of the United States, women are more likely to have adverse events from vaccination with this vaccine, which was supported by the results of this study.

The incidence of adverse events in this study was higher than that reported in previous reports of adverse events from the COVID-19 Vaccine Moderna Intramuscular Injection in Japan and other countries^1,3,4)^.

We considered that this reason is because this study group included more younger women than those in previous studies.

In reports performed in foreign countries, headache and fatigue were the most frequently reported compared to other systemic events ^3)^. However, in this study, many cases of myalgia and fatigue were also observed. In this study, we found that female sex was a risk factor for myalgia. This study is characterized by a higher proportion of younger people and women than those of other reports, which was also reflected in the incidence of adverse events ^3)^.

In this study, to obtain information of adverse events that occurred after leaving the site, we conducted a questionnaire survey on a website. Therefore, the existence of a reporting bias cannot be completely ruled out. This is one of the limitations of the present study.

Information on adverse events that did not require treatment was not available, except for voluntary reports. This is a general limitation in adverse event investigations.

The COVID-19 Vaccine Moderna Intramuscular Injection efficacy after second doses was 94.1% in preventing symptomatic, laboratory-confirmed COVID-19 among persons without evidence of previous SARS-CoV-2 infection ^3,5)^. This vaccine requires two times injections to be fully effective, with the second dose having a higher frequency of adverse events than the first time injection^1-4)^. Although there are no detailed data on Japanese who received the COVID-19 Vaccine Moderna Intramuscular Injection, data on Japanese healthcare workers who received the COMIRNATY intramuscular injection show that female and young people are more likely to increase of adverse events at the time of the second dose ^2)^. Similar adverse events are predicted with the COVID-19 Vaccine Moderna Intramuscular Injection. In addition, there is a high possibility that this vaccine will be administered in younger people. Age is often a risk factor for adverse events and should be pursued with caution. However, the adverse events caused by vaccination with this vaccine are rarely serious, and the benefits of vaccination should be emphasized ^6)^. For peace of mind at the time of vaccination, those with many risk factors or those who had adverse events at the time of the first inoculation should prepare antipyretic analgesics, such as acetaminophen, before the second inoculation in anticipation of any adverse events. We will also investigate, analyze, and report adverse events after the second vaccination. This information will give impacts on considering adverse events and its mechanisms in mRNA vaccination.

## Data Availability

Due to the nature of this research, participants of this study did not agree for their data to be shared publicly, so supporting data is not available.

